# Validation of a Self-administrable, Saliva-based RT-qPCR Test Detecting SARS-CoV-2

**DOI:** 10.1101/2020.06.05.20122721

**Authors:** Meghan Miller, Michael Jansen, Alexander Bisignano, Shannon Mahoney, Christine Wechsberg, Nikita Albanese, Louise Castillo, Pia Farinas, Gabriel A. Lazarin, Malgorzata Jaremko

**Affiliations:** Phosphorus Diagnostics, New York, NY, USA.

## Abstract

**Background:** Rapid and easy COVID-19 diagnostic testing is essential to controlling the pandemic and facilitating safe resumption of clinical care, employment and other social activities.

**Methods:** This study was conducted to validate an self-administrable saliva-based RT-qPCR test for the SARS-CoV2 virus under controlled laboratory conditions (analytical validation) according to federal guidance. An additional clinical study assessed positive (n = 34) and negative (n = 57) nasopharyngeal swab samples collected contemporaneously with saliva samples. Assessments for analytical specificity, sensitivity, cross reactivity and sample stability to simulate shipping conditions were conducted.

**Results:** Positive and negative agreement with third-party laboratory results were reported as 97.1% and 96.5–98.2%, respectively. Limit of detection was established at 5 copies/µL. Stability through simulated shipping conditions found 100% concordance up to 56 hours after collection. Discussion: These data validate a self-collected saliva-based COVID-19 RT-qPCR assay that performs comparably well to an assay of health care-provider administered nasopharyngeal swab samples. Accordingly, the United States Food and Drug Administration granted emergency use authorization in June 2020. Use of the saliva-based assay overcomes barriers to the necessary widespread testing, including strained health care resources, supply chain disruptions of laboratory materials, testing and protective equipment and exposure risks due to close interpersonal contact.

## Introduction

COVID-19, caused by the SARS-CoV-2 virus, manifests a range of nonspecific symptoms, from asymptomatic to respiratory distress, fever, cough, severe pneumonia, multi-organ involvement and death. At the time of publication, the United States recorded more than 1,700,000 cases of COVID-19, over 100,000 deaths and, with nearly 6,000,000 diagnosed worldwide, the disease is designated a pandemic by the World Health Organization[1,2]. Rapid and vast spread is partly attributed to asymptomatic patients being contagious for indeterminate time periods. Additional contributions to spread include the various modes of infection including person-to-person contact, contact with contaminated surfaces and objects, and through respiratory droplets[3–5].

Effective tracing and containment of COVID-19 requires widespread testing, which to date has been limited in capacity and accessibility. Testing is primarily performed on a nasopharyngeal swab, which requires administration by trained health care professionals.[6,7]. This combination of substantial resource usage and limited supply of testing has limited the ability to diagnose and manage individuals.

Self-administered saliva collection potentially overcomes barriers to widespread testing by removing the need for sample collection by trained healthcare professionals and decreases the exposure and virus transmission risk associated with current testing methods. Early studies demonstrated 91.7% – 100.0% concordance in 12 and 25 patients with virus detected by naso- or oro-pharyngeal swabs[8,9]. Our study evaluated performance of saliva specimens obtained from a self-administered collection tube for detection of SARS-CoV2, in comparison with current nasopharyngeal swab testing methods that have received an Emergency Use Authorization (EUA) from the Food and Drug Administration (FDA).

## Methods

This study describes analytical and clinical validation of saliva collection for SARS-CoV2 detection by RT-qPCR. Paired clinical samples, self-collected saliva and clinician-administered nasopharyngeal swabs (NPS), were contemporaneously collected and results were compared.

This study was conducted with review and approval of the Aspire Institutional Review Board (Protocol #20200726).

### Testing Methods

All testing was performed at Phosphorus Diagnostics, certified under the Clinical Laboratory Amendments (31D2123554), accredited by the College of American Pathologists (9504988) and licensed by New York (PFI 9267) and New Jersey (0010873).

The COVID-19 RT-PCR Test is a real-time reverse transcription polymerase chain reaction (rRT-PCR) test. The test uses two primers and probes sets to detect two regions in the SARS-CoV-2 nucleocapsid (*N*) gene (N1 and N2) and one primer and probe set to detect human RNase P (RP) in a clinical sample.

Collected saliva was received at the laboratory and incubated for one hour at 50℃ to inactivate RNase present in the specimen. Paired NPS were collected using and received at the laboratory at the same time. Three RNA extraction methods were evaluated: manual methods using the MagMAX™ Viral/Pathogen Nucleic Acid Isolation Kit (ThermoFisher Scientific) or Maxwell® HT Viral TNA Kit (Promega Corporation) and automated extraction using the Maxwell RSC TNA Viral Kit (Promega Corporation). Extracted RNA was reverse transcribed to cDNA and subsequently amplified using the CFX384 Touch Real-Time PCR Detection System with CFX Manager software version 3.1 (Bio-Rad Laboratories).

The PCR conditions including thermocycler program, reaction mix, and Ct threshold were first established using positive (2019-nCoV_N_Positive Control, Integrated DNA Technologies) and negative controls, and further tested on RNA extracted from patients’ saliva. During the amplification process, the probe anneals to a specific target sequence located between the forward and reverse primers. During the extension phase of the PCR cycle, the 5’ nuclease activity of Taq polymerase degrades the bound probe, causing the reporter dye (FAM) to separate from the quencher dye (BHQ1), generating a fluorescent signal.

Quality controls on every assay included a negative no template control to rule out sample contamination, a positive template control, an internal control targeting RNase P to verify presence of nucleic acid and a negative extraction control to monitor for sample cross contamination.

Analysis of the fluorescence curves generated during the PCR process is done using the Bio-Rad CFX Manager software version 3.1. The threshold is set during the exponential phase of the PCR run. This exponential phase is defined as the time at which the PCR product begins to amplify, with the amount of product doubling after every cycle. The cycle number that each sample’s amplification curve passes this threshold is known as the Ct value. This value is what is used by the end user to determine the presence of viral RNA in the initial sample.

Results were interpreted according to the findings described in Table 1. If any or all of the negative no template, positive or internal controls did not meet acceptability criteria, the RT-PCR run was repeated. If the negative extraction control did not meet acceptability criteria, all specimens on the bath were re-extracted and re-run on RT-PCR.

**Table 1.** Results interpretations.

| **N1** | **N2** | **RP** | **Results Interpretation** |
| --- | --- | --- | --- |
| + | + | +/- | SARS-CoV-2 detected |
| If any of the two targets is positive |  | +/- | Inconclusive results |
| - | - | + | SARS-CoV-2 not detected |
| - | - | - | Invalid Results |

N1, N2 refer to two regions of the SARS-CoV-2 nucleocapsid (*N*) gene. RP refers to human RNase P.

### Clinical Population

Eligible individuals were identified by referral from physicians at two primary care medicine facilities in New York City. Individuals were eligible or ineligible according to the criteria described in Table 1.

Participants’ clinical findings (symptoms at onset, date of onset, current symptoms, medication history, pre-existing health conditions) were recorded. Samples from nasopharyngeal swabs (NPS) and saliva were collected within the same clinical visit. Clinical trial coordinators performed NPS collections and observed saliva collections. Saliva was collected using the Orasure Oragene®·Dx (OGD-510) device according to manufacturer instructions.

Paired samples were transported at ambient temperature and tested at Phosphorus within 48 hours after collection. NPS were placed in a viral transport medium.

#### Data Entry

Upon arrival to the laboratory, the samples were de-identified and assigned a unique barcode before laboratory testing began. Designated personnel entered patient information submitted on the requisition form together with unique barcodes assigned to the patient into the password protected and HIPAA-compliant file document, which served as a key. The uniquely barcoded sample was entered into the Phosphorus laboratory information system, Elements, and laboratory testing personnel remained blinded to the patient’s prior

RT-qPCR results, or any other clinical information. Upon completion of the wet bench analysis, a designated personnel provided a summary of comparison of prior COVID-19 testing results with those received by Phosphorus laboratory. At that time, study personnel were granted access to other clinical information entered into the key file for purposes of further analysis.

## Results

### Analytical Validation

#### Limit of Detection and Analytical Sensitivity

Preliminary analysis was based on synthetic SARS-CoV-2 RNA (Twist Bioscience) diluted to 10,000 copies/µL and then spiked into previously negative self-collected saliva samples concentrations of 1000, 500, 200, 100 and 5 copies/µL in triplicate. The triplicates were extracted as described above. Each sample was run on RT-qPCR and based on this, the limit of detection (LoD) was preliminarily established at 5 copies/µL for all extraction methods.

To confirm this LoD, 20 individual replicates of previously negative saliva samples were spiked with the same synthetic SARS-CoV-2 RNA at 5 copies/µL and underwent the previously described extraction methods. RT-qPCR analysis on detected 100% of N1 and N2 targets for the two manual extraction methods (20/20 and 20/20) and 95% of N1 (19/20) and 100% (20/20) N2 targets using the automated extraction method. Table 3 summarizes these data.

**Table 2.** Inclusion and exclusion criteria for clinical population.

| Inclusion criteria | Exclusion criteria |
| --- | --- |
| ● 18 years of age or older, and<br>● Reporting any of the coronavirus suggesting symptoms including but not limited to respiratory symptoms, cough, sore throat, headache, fever, asthenia/fatigue/ malaise, myalgia/aches, chills/sweating, diarrhea, dyspnea, pneumonia, respiratory distress, multi-organ involvement, and<br>○ Having positive RT-qPCR test performed on nasopharyngeal swab within <5 days, or<br>○ Undergoing RT-qPCR testing on nasopharyngeal swab at the time of enrollment<br>● Or, asymptomatic patient with RT-qPCR test results performed on nasopharyngeal swab with positive or negative results within of research specimen collection | Individuals were excluded from participation if they met any of the following criteria:<br>● Not having RT-qPCR test performed on nasopharyngeal swab previously, or not undergoing such testing at the time of enrollment<br>● Confirmed infection with any other respiratory virus or bacteria<br>● History of non-infectious diseases such as vasculitis, dermatomyositis, idiopathic pulmonary fibrosis and organizing pneumonia<br>● Less than 18 years of age<br>● Currently pregnant |

**Table 3.** Summary of the limit of detection verification study for 20 replicates at 5 copies/ µL using three extraction methods.

| RNA Extraction Method | Detected Mean Ct (Standard Deviation) |  |  | Detection Rate (n, Detected / n, Total) |  |  |
| --- | --- | --- | --- | --- | --- | --- |
|  | N1 | N2 | RP | N1 | N2 | RP |
| MagMAX Viral/Pathogen Nucleic Acid Isolation Kit | 34.87<br>(0.7) | 35.50<br>(0.7) | 23.40<br>(0.5) | 100%<br>(20/20) | 100%<br>(20/20) | 100%<br>(20/20) |
| Maxwell HT Viral TNA Kit | 35.36<br>(0.9) | 36.89<br>(0.9) | 22.52<br>(0.4) | 100%<br>(20/20) | 100%<br>(20/20) | 100%<br>(20/20) |
| Maxwell RSC TNA Viral Kit | 35.57<br>(1) | 36.95<br>(1.2) | 22.58<br>(0.7) | 95%<br>(19/20) | 100%<br>(20/20) | 100%<br>(20/20) |

#### Saliva Sample Stability

In order to ensure integrity of the sample after self collection and transport, sample stability over time was studied. Forty self-collected saliva samples were assessed, including “high positives” (n = 10 spiked at 25–50 copies/µL), “low positives” (n = 20 spiked at 10 copies/µL) and negatives (n = 10). Conditions simulated a local storage after collection at high temperature (40℃) for 8 hours and 48-hour shipping cycle (22℃ to 40℃), for a total of 56 hours from collection to commencement of analysis. After separation into triplicates and extraction according to the three aforementioned protocols, tested samples yielded 100% agreement with expected results.

#### Cross-reactivity and Analytical Specificity

This test utilizes the 2019-nCoV Centers for Disease Control and Prevention (CDC) EUA Authorized qPCR Probe Assay primer/probe mix. *In silico* BLASTn analysis of primer and probe specificity was performed by the CDC[10].

Additionally, we performed wet bench testing of the plasmid controls for Middle East respiratory syndrome and severe acute respiratory syndrome (MERS_CoV control and SARS_CoV control, Integrated DNA Technologies). All results were negative.

### Clinical Validation

To analyze the clinical performance, results from self-collected saliva were compared to those of nasopharyngeal swab (NPS), collected within the same visit at one of two New York City physician offices.

Paired samples were tested after three aforementioned extraction methods. In total 34 NPS positive and 57 NPS negative samples were tested for each (Table 4). Positive agreement was 97.1% (CI: 85.1–99.5%) for each method. Negative agreement was 98.2% (CI: 90.7–99.7%) using Maxwell HT Viral TNA Kit extraction and 96.5% (CI: 88.1–99.0%) using the remaining two methods.

**Table 4.** Results from paired self-collected saliva and nasopharyngeal swab in COVID-19 patients, using three extraction methods, using NPS as the reference and stratified by extraction method.

| NPS Result (n, patients) | Sample Type | Analysis | Target |  |  |
| --- | --- | --- | --- | --- | --- |
|  |  |  | N1 | N2 | RP |
| MagMAX Viral/Pathogen Nucleic Acid Isolation Kit |  |  |  |  |  |
| NPS Positive (n=34) | NPS | Positive (%) | 34 (100%) | 34 (100%) | 34 (100%) |
|  |  | Ct, mean (SD) | 34.5 (5.0) | 35.4 (5.2) | 22.6 (2.0) |
|  | Saliva | Positive (%) | 33 (97.1%) | 33 (97.1%) | 33 (97.1%) |
|  |  | Ct, mean (SD) | 33.2 (3.9) | 34.4 (4.0) | 23.1 (2.5) |
| NPS Negative (n=57) | NPS | Positive (%) | 0 (0%) | 0 (0%) | 57 (100%) |
|  |  | Ct, mean (SD) | N/A | N/A | 26.5 (2.8) |
|  | Saliva | Positive (%) | 2 (3.5%) | 2 (3.5%) | 55 (96.5%) |
|  |  | Ct, mean (SD) | 34.5 (3.5) | 35.3 (2.6) | 21.7 (1.0) |
| Promega Maxwell HT Viral TNA Kit |  |  |  |  |  |
| NPS Positive (n=34) | NPS | Positive (%) | 34 (100%) | 34 (100%) | 34 (100%) |
|  |  | Ct, mean (SD) | 34.8 (4.5) | 35.7 (4.6) | 24.0 (1.6) |
|  | Saliva | Positive (%) | 33 (97.1%) | 33 (97.1%) | 33 (97.1%) |
|  |  | Ct, mean (SD) | 34.0 (2.7) | 35.5 (2.9) | 23.6 (1.9) |
| NPS Negative (n=57) | NPS | Positive (%) | 0 (0%) | 0 (0%) | 57 (100%) |
|  |  | Ct, mean (SD) | N/A | N/A | 27.9 (2.6) |
|  | Saliva | Positive (%) | 1 (1.8%) | 1 (1.8%) | 56 (98.2%) |
|  |  | Ct, mean (SD) | 32.6 (0) | 33.6 (0) | 23.0 (1.0) |
| Maxwell RSC TNA Viral Kit run on Maxwell RSC 48 System |  |  |  |  |  |
| NPS Positive (n=34) | NPS | Positive (%) | 34 (100%) | 34 (100%) | 34 (100%) |
|  |  | Ct, mean (SD) | 34.6 (4.9) | 35.5 (5.1) | 22.8 (1.5) |
|  | Saliva | Positive (%) | 33 (97.1%) | 33 (97.1%) | 33 (97.1%) |
|  |  | Ct, mean (SD) | 34.1 (2.6) | 35.5 (2.7) | 23.1 (1.9) |
| NPS Negative (n=57) | NPS | Positive (%) | 0 (0%) | 0 (0%) | 57 (100%) |
|  |  | Ct, mean (SD) | N/A | N/A | 26.7 (2.5) |
|  | Saliva | Positive (%) | 2 (3.5) | 2 (3.5) | 55 (96.5) |
|  |  | Ct, mean (SD) | 35.0 (2.7) | 36.7 (2.7) | 22.1 (1.1) |

\*NPS = nasopharyngeal swab, SD = standard deviation

## Discussion

This study validates the performance of a self-collected saliva-based RT-qPCR test to detect SARS-CoV-2 in symptomatic patients. The FDA granted emergency use authorization for this test in June 2020 [cite]. To our knowledge, this is the first published peer-reviewed study of a clinically available and FDA authorized test.

Three RNA extraction methods were evaluated yielded a sensitivity of 97.1% sensitivity and 96.5–98.2% specificity relative to nasopharyngeal swab results.

This study builds upon limited data that demonstrate comparable performance of saliva in comparison to nasopharyngeal swab. Peer-reviewed studies are limited in number and scale. A study of 33 tests in 25 COVID-19 patients in Italy found 100% positive agreement after diagnosis by nasopharyngeal swab[9]. In fact, saliva samples remained positive for a longer period of time than respiratory samples, suggesting the possibility that patients may be contagious by saliva for a greater length of time than what is identifiable on a respiratory sample. The utilization of this collection method may provide more informative information about recovery and evolution of the virus than oropharyngeal or nasopharyngeal swabs[9].

Another Hong Kong-based study found 91.7% concordance among saliva and nasopharyngeal swab samples in a population of 12 patients. Viral load was monitored regularly in this population and found decreasing levels in saliva over time[8]. Studies available in pre-print describe findings generally consistent with the ones described here and our data[11–13], and another concludes saliva to be an inferior sample, though the study design includes confounding factors and inconsistencies in study design that could benefit from peer review[14].

Several considerations led to the use of saliva and three extraction methods. Current supply chain issues of swabs, viral transport medium, personal protective equipment and reagents have hindered COVID-19 viral test development and widespread usage. This approach using an alternative sample type validated using multiple extraction methods mitigates the risk of disruptions.

A saliva collection method also minimizes exposure risks because it can be self-administered in or out of clinical settings.

This validation of a novel testing device for COVID-19 serves to increase access to vital testing to a broader population. Access to COVID-19 testing for frontline healthcare workers, essential employees and general members of the population, both symptomatic and asymptomatic, will ultimately reduce the spread of disease and decrease the burden on the healthcare system.

## Data Availability

Data is publically available by contacting authors or the FDA.

## Acknowledgments

The authors thank the dedicated research coordinators, Amanda Berg, Sebastian Bidegain, Dana Golden and Cara Stefanacci for their contributions to this project. Additionally, the authors thank the study sites that enabled this research to be conducted by granting access to their patients.

